# Developing Provider-Co-Created Prototypes Addressing Equity-Related Barriers in Liver Transplantation for Hepatocellular Carcinoma

**DOI:** 10.64898/2026.05.15.26353301

**Authors:** Lauren D. Nephew, Courtney Moore, Nicole Garcia, Lisa Parks, Allison McKay, Sebastian Abad, Susan M. Rawl

**Affiliations:** Division of Gastroenterology and Hepatology, Department of Medicine, Indiana University, Indianapolis, IN, USA; Research Jam, Community Health Partnerships, Indiana Clinical and Translational Sciences Institute, Indiana University School of Medicine, Indianapolis, IN, USA; Indiana University School of Nursing, Indiana University Simon Cancer Center, Indianapolis, IN, USA

**Keywords:** Disparities, cirrhosis, social determinants of health, barriers, interventions

## Abstract

**Background:** Black patients and individuals with low socioeconomic status (SES) face significant disparities in accessing curative therapies for hepatocellular carcinoma (HCC), including liver transplantation. This study aimed to develop provider-co-created intervention prototypes in response to patient-identified barriers and recommendations.

**Methods:** A human-centered design session with hepatology and transplant providers at a large academic medical center was conducted. Prior to the session, participants were presented with barriers and preliminary solutions identified through an earlier human-centered design session with Black and low-SES patients. Using structured ideation methods, including brainwriting, challenge mapping, and concept voting, providers co-created intervention prototypes. Final concepts were synthesized from patient insights, provider input, and design methods using affinity diagramming and concept modeling.

**Results:** Nine providers participated in the session. They focused on three key areas for intervention: inefficiencies in transplant pre-evaluation, inadequate social support, and information overload. Solutions included: (1) a structured triage pathway to standardize referrals and reduce delays; (2) a peer navigator model to guide patients through the transplant process; and (3) a multimodal transplant education roadmap to improve comprehension and engagement. These prototypes addressed both patient- and system-level barriers.

**Conclusions:** Protypes developed through provider-led design, grounded in patient-identified barriers and co-created ideas, can yield actionable, scalable strategies to advance equity in HCC care. Future work will refine these prototypes through patient feedback and pilot them in clinical settings.

## INTRODUCTION

In the United States, hepatocellular carcinoma (HCC) remains one of the fastest-rising causes of cancer-related death, driven by rising rates of cirrhosis, persistent racial and socioeconomic disparities, and delayed access to curative treatment^1,2^. For early-stage disease, curative therapies such as liver resection and liver transplantation (LT) provide the best long-term outcomes^3^. However, access to these treatments remains inequitable, disproportionately affecting Black patients and individuals experiencing adverse social determinants of health (SDOH)^4^. These disparities contribute to worse survival, underscoring the urgent need for interventions that address systemic barriers to care^5,6^.

Reducing disparities in access to LT requires a multidisciplinary, patient-centered approach. However, systemic barriers persist throughout the transplant process. Fragmented communication among providers, logistical complexities during evaluation, and misaligned care coordination efforts often lead to unnecessary delays and missed opportunities for curative treatment^7,8^. While research underscores the value of culturally sensitive, patient-centered interventions, particularly for socioeconomically disadvantaged populations, efforts to address disparities in LT lag behind those in kidney transplantation (KT), despite similar needs.

Interventions implemented in KT such as telehealth services, structured patient education, and coordinated care models have shown promise in reducing disparities and improving access^9,10^. For example, telemedicine has facilitated pre-evaluation visits and reduced logistical barriers, while peer navigation programs have improved engagement among marginalized patients^11^. The COVID-19 pandemic further accelerated the adoption of virtual evaluation models in solid organ transplantation, helping maintain access to care during disruptions^12^. While similar strategies have been explored in LT, their adoption has been more limited and inconsistent. Moreover, despite growing evidence supporting stakeholder-driven approaches like human-centered design (HCD) in other healthcare settings, these methods remain underutilized in transplant intervention development, particularly in LT.

This study aims to address gaps in equity-focused interventions in LT by leveraging a human-centered approach to develop early-phase, provider-informed, patient-centered intervention prototypes. Building on prior research identifying patient-reported barriers to curative therapy, this study engages physicians, social workers, and care coordinators to design practical, scalable interventions that improve care coordination, patient communication, and access to life-saving therapies^13^. By integrating insights from both patient and provider perspectives, this work seeks to bridge the gap between disparities research and actionable solutions, ensuring that interventions are aligned with real-world clinical practice and the needs of underserved populations.

## METHODS

This study employed human-centered design (HCD) methodology to develop solutions addressing patient-identified barriers to curative therapy for hepatocellular carcinoma (HCC)^14^. HCD actively engages stakeholders throughout the design process to ensure feasible, patient-centered solutions. This work builds on prior HCD sessions with Black and low socioeconomic status (SES) patients that identified barriers to HCC treatment, including distrust in the healthcare system, difficulties navigating transplant, information overload, fragmented communication, limited social support, travel burden, and financial hardship^13^. Patients also proposed potential solutions such as clearer educational materials, progress updates, and peer or partner support^13^. This study was approved by the first author’s Institutional Review Board and conducted in accordance with the Declaration of Helsinki.

### Setting

Unlike traditional focus groups, HCD sessions use interactive activities to surface explicit and latent challenges and foster iterative feedback, yielding interventions adaptable to clinical settings ^14^. This study was conducted in a virtual format to accommodate participants’ demanding schedules. Sessions were facilitated by an HCD team housed within the Clinical and Translational Sciences Institute (CTSI). Researchers engaged healthcare professionals involved in hepatology and transplant care to gather insights on improving access to curative therapies for HCC patients. Participants were selected for their clinical experience managing HCC and LT patients.

### Sampling

Eligible participants included physicians in hepatology, transplant medicine, oncology, interventional radiology, and general surgery, as well as advanced practice providers, transplant nurse coordinators, and transplant social workers. Recruitment occurred via email and phone within a large Midwestern academic health center. Sixteen providers consented, and nine attended: five physicians, three registered nurses, and one transplant social worker.

### Information Collection

Patient-identified barriers were translated into “How Might We” (HMW) questions to guide provider sessions. These focused on three challenge areas: improving the transplant pre-evaluation process, enhancing social support, and addressing information-related challenges. Framing discussions in this way ensured that provider-generated solutions remained grounded in patient priorities.

Discussions were conducted via Zoom, with Miro, an online whiteboarding tool, used for structured brainstorming and collaborative problem-solving. To facilitate open discussion and encourage equal participation, the session was divided into two breakout groups: one for physicians and another for coordinators, social workers, and other care providers. This separation allowed each group to explore challenges from their unique perspectives without hierarchical dynamics influencing the conversation. Following the breakout sessions, participants reconvened to share their insights and synthesize key takeaways.

### Ideation Process and Prototype Development

Each session followed a structured, facilitator-led format, ensuring that discussions remained focused, participatory, and solution-oriented. The integration of digital tools enabled real-time engagement, allowing providers to refine interventions collaboratively. Sessions began with an overview of patient-identified challenges. Physicians addressed “How might the transplant pre-evaluation process be improved?” while coordinators, social workers, and other providers focused on “How might we help patients have better support?” Participants then used brainwriting to generate ideas independently before group discussion, clustering, and refinement into feasible interventions. A “switch and vote” exercise allowed groups to review and build on each other’s ideas, then vote on the most promising concepts based on feasibility, patient impact, and alignment with transplant system constraints.

The research team facilitated a concept refinement phase where top-ranked solutions were further developed through iterative discussion and structured feedback. This ensured proposed interventions were practical, scalable, and responsive to patient-identified challenges. Final prototypes were made visual using diagramming tools in Miro as participants described their concepts. These prototypes formed the foundation for the next phase of refinement and patient-centered evaluation. At the close of the session, all participants reconvened to apply brainwriting to information-related challenges, generating multiple ideas per prompt to expand the solution set.

### Data Analysis

The primary goal of the analysis was to identify dominant themes and synthesize insights from provider discussions. Affinity diagramming, a qualitative analysis technique, was used to organize and interpret data. All sticky notes generated during brainstorming were captured in Miro, eliminating the need for extensive pre-processing. The research team reviewed and grouped related ideas into three major challenge areas: transplant pre-evaluation, social support, and information barriers.

Overlapping themes were consolidated into a comprehensive framework and layered onto a flow diagram to visualize how provider-generated solutions mapped onto the transplant evaluation process. This approach ensured solutions were directly linked to patient-identified barriers.

Synthesis focused on moving from “what is” to “what could be,” turning key themes into strategies. The research team assessed the final affinity diagram alongside concepts from the last brainstorming round, identifying four major strategies based on feasibility, relevance, and potential impact on transplant evaluation, communication, and social support. Findings were categorized into intervention-oriented frameworks aligned with study objectives.

## FINDINGS

The creation session had nine participants: five physicians (four transplant hepatologists and one interventional radiologist), three RN transplant coordinators, and one transplant social worker.

### Thematic Analysis of Key Challenges & Potential Solutions

Nine major themes were identified that aligned with three primary challenge areas explored in the facilitated discussions: (1) the transplant pre-evaluation process, (2) gaps in social support, and (3) information challenges. These themes represent provider-identified barriers and conceptual solutions, reflecting their perspectives on addressing challenges raised by Black and low SES patients with HCC.

*Transplant Pre-Evaluation Process: How might the transplant pre-evaluation process be improved?*

Providers identified multiple barriers within the transplant pre-evaluation process, highlighting inefficiencies in referral patterns, evaluation logistics, and provider communication. A key challenge was the <u>inappropriate or delayed referral of patients</u>, leading to prolonged wait times and inefficient use of limited evaluation slots. As one provider noted, “*New patient spots are very limited. The number of inappropriate referrals we receive is a lot, and it would help eliminate no-shows*” (Coordinators & Social Worker). To address this, participants proposed a triage pathway where an experienced pre-transplant coordinator would conduct pre-screening to determine whether a patient required a full transplant evaluation or should be directed to general hepatology. Some also recommended the use of virtual pre-evaluation clinics, where a physician could assess borderline or unclear cases without requiring a full in-person evaluation.

In addition to referral inefficiencies, providers described <u>logistical delays in the pre-evaluation process</u>, often requiring multiple visits for laboratory testing, imaging, and psychosocial assessment. Many advocated for a “transplant evaluation sprint” model that would consolidate all required testing into a single visit spanning two to three days rather than spreading these assessments over multiple appointments. “*Bring patients in and do all testing in 3 days—block slots out for patients so the entire process is streamlined*” (Coordinators & social workers).

In addition to process inefficiencies, providers cited <u>poor communication</u> between transplant teams, hepatologists, oncologists, and social workers as a major bottleneck. Much of their time was spent tracking down missing records, making phone calls, and waiting for faxed documents rather than focusing on patient care. One provider explained, “*All of us deal with HCC patients, I see more of them because of the doctor I work with in transplant. There is some communication between us and the physicians who are treating the cancer. But I definitely feel like it could be improved upon*.” (Coordinators & Social Work). Standardizing communication through automatic data-sharing systems was proposed as an intervention to improve care coordination and ensure that all providers were aligned in their decision-making. Together, these findings underscore the need for structured referral pathways, more efficient evaluation processes, and improved provider communication to streamline transplant pre-evaluation and optimize patient outcomes.

#### Social Support

Providers highlighted significant gaps in support for patients and caregivers throughout the transplant process, emphasizing the need for structured guidance, clear expectations, and additional staff to reduce provider workload.

Many patients lacked <u>instrumental support</u>, such as transportation assistance, financial resources, and engaged caregivers, making it difficult to navigate the complex transplant journey. As one provider noted, “*It would be awesome if patients had a navigator, like with cancer treatment at cancer centers. Someone other than a coordinator that has so many demands*” (Coordinators and Social Worder) and another that “*Worth remembering that the caregivers of patients with organ failure themselves need suppor*t” (Physicians). To address this, participants proposed implementing a peer navigator program modeled after cancer patient navigation programs, where trained individuals, including former transplant recipients, could assist patients in coordinating care, understanding expectations, and accessing available resources.

Beyond logistical and emotional support, providers also underscored the importance of setting <u>realistic expectations</u> upfront, particularly regarding transplant eligibility and post-surgical recovery. Many patients were unaware of the stringent requirements for transplant candidacy, which led to frustration and delays when they were deemed ineligible late in the process. “*If patients understood the broad requirements for transplant at the very beginning of the process, it might be clear sooner that they will not be accepted*” (Physicians), one provider explained. To mitigate these challenges, providers recommended standardizing eligibility communication by offering a “What to Expect” curriculum, which could include structured education sessions, written guidelines, and pre-consultation checklists to ensure patients fully understood transplant prerequisites and milestones.

Additionally, providers expressed concerns that <u>high patient volumes</u> and staffing limitations were hindering individualized support. Many coordinators and social workers were overwhelmed with large caseloads, limiting their ability to provide personalized guidance and follow-up care. One provider noted, “*My pre-transplant coordinator rarely works less than 50 hours a week*.” (Physicians). To alleviate this burden, participants advocated for hiring additional coordinators, particularly a dedicated HCC transplant coordinator, to address the unique needs of liver cancer patients. Expanding the care team to include lay navigators and non-clinical staff was also proposed as a way to shift logistical and administrative responsibilities away from clinical personnel, allowing them to focus on direct patient care. These findings highlight the urgent need for increased support structures, effective patient education, and expanded staffing to enhance the transplant experience and improve outcomes.

#### Information Challenges

Providers highlighted significant <u>gaps in patient education</u>, emphasizing the need for structured, multi-format tools to help patients better understand their diagnosis, treatment options, and the transplant process. Many patients struggled to navigate complex medical information, particularly regarding insurance, eligibility, and long-term post-transplant care. To address this, providers recommended expanding educational resources to include pre-recorded videos, in-person education sessions, and visual aids such as charts, infographics, and checklists. One provider from the Coordinator and Social Worker group noted w*e need regularly occurring information sessions that could be accessed online and on-demand—patients need access to structured, digestible education* (Coordinators and Social Worker). These resources would allow patients to access information at their own pace while reinforcing key messages over time.

In addition to improving how information is delivered, providers stressed the importance of summarizing key takeaways after medical visits to prevent <u>information overload</u>. Many patients struggled to retain details from consultations, especially after receiving a cancer diagnosis, leading to confusion about next steps. One provider explained, “*The model we have now is we just dump all this information in one go and its very overwhelming and they don’t remember it, and sometimes they are embarrassed to ask like, ‘I know you told me this*.’” (Coordinators and Social Workers). To address this, providers proposed creating structured visit summaries that included a treatment roadmap, calendar-based appointment tracking, and a digital portal where patients could review test results and upcoming milestones in real time.

Despite consensus on the need for better educational tools, providers expressed frustration that limited institutional support and funding hindered the implementation of these interventions. Many proposed solutions they felt would remain underfunded or logistically unfeasible given current resources. One participant described the strain on existing systems, stating, “*What you need is enough money or support to facilitate the patient experience” (Physicians). “Right now, we have more patients coming in than going out—not enough space or resources to keep up*” (Coordinators, Social Workers). To bridge this gap, providers recommended advocating for institutional investment in dedicated HCC support programs, including funding for housing assistance, transportation services, and patient navigation teams.

Together, these insights highlight the need for comprehensive, accessible, and well-resourced educational strategies to improve patient understanding, retention, and engagement in their transplant journey.

### From Barriers to Provider-Generated Prototypes

#### Provider-Generated Prototypes for Addressing Transplant Barriers

Through structured ideation, providers developed several concepts to address patient- and system-level challenges in the transplant evaluation process.

The Transplant Triage Pathway aimed to standardize referral guidelines for hepatologists and streamline pre-evaluation. A virtual clinic was proposed for initial assessments, psychosocial screening, and eligibility review, reducing inefficiencies and optimizing clinic availability (Figure 1).

**Figure 1.**
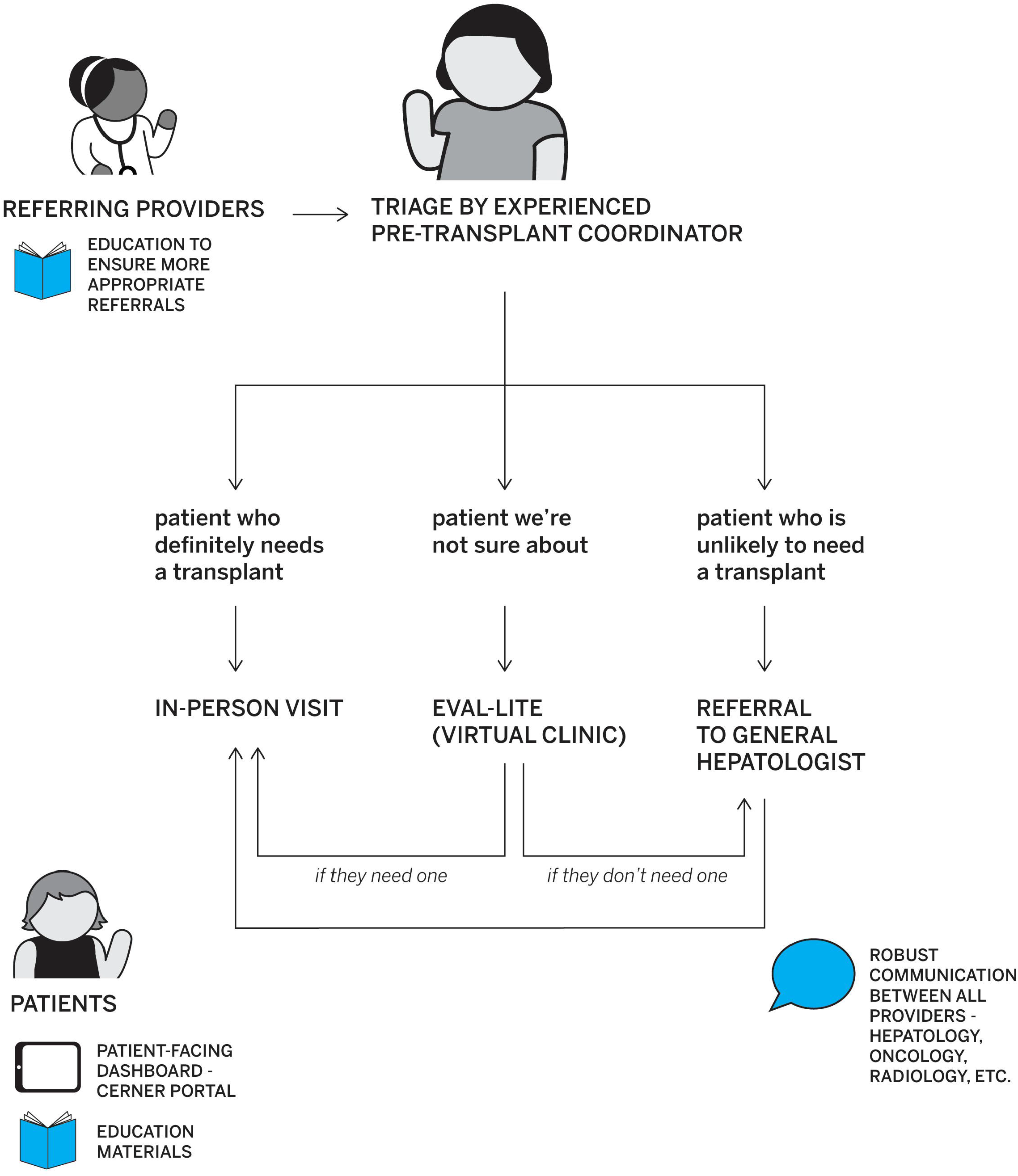
Provider-Generated Prototype:Transplant Triage Pathway. Structured triage pathway to standardize transplant referrals and reduce inappropriate or delayed evaluations. An experienced pre-transplant coordinator conducts an initial triage to determine whether patients require a full evaluation, can be directed to general hepatology, or can be monitored with a limited “Eval-Lite” virtual clinic. The model incorporates robust provider communication across specialties, patient-facing dashboards and education materials, and referral feedback to improve appropriateness of transplant referrals.

To address information overload, providers designed a Roadmap for Transplant Education, a stage-based framework delivering consistent information at key milestones. Components included a visual roadmap of HCC treatment, appointment and testing checklists, multimedia tools, and a digital progress tracker. This prototype sought to improve comprehension, reduce confusion, and standardize messaging (Figure 2).

**Figure 2.**
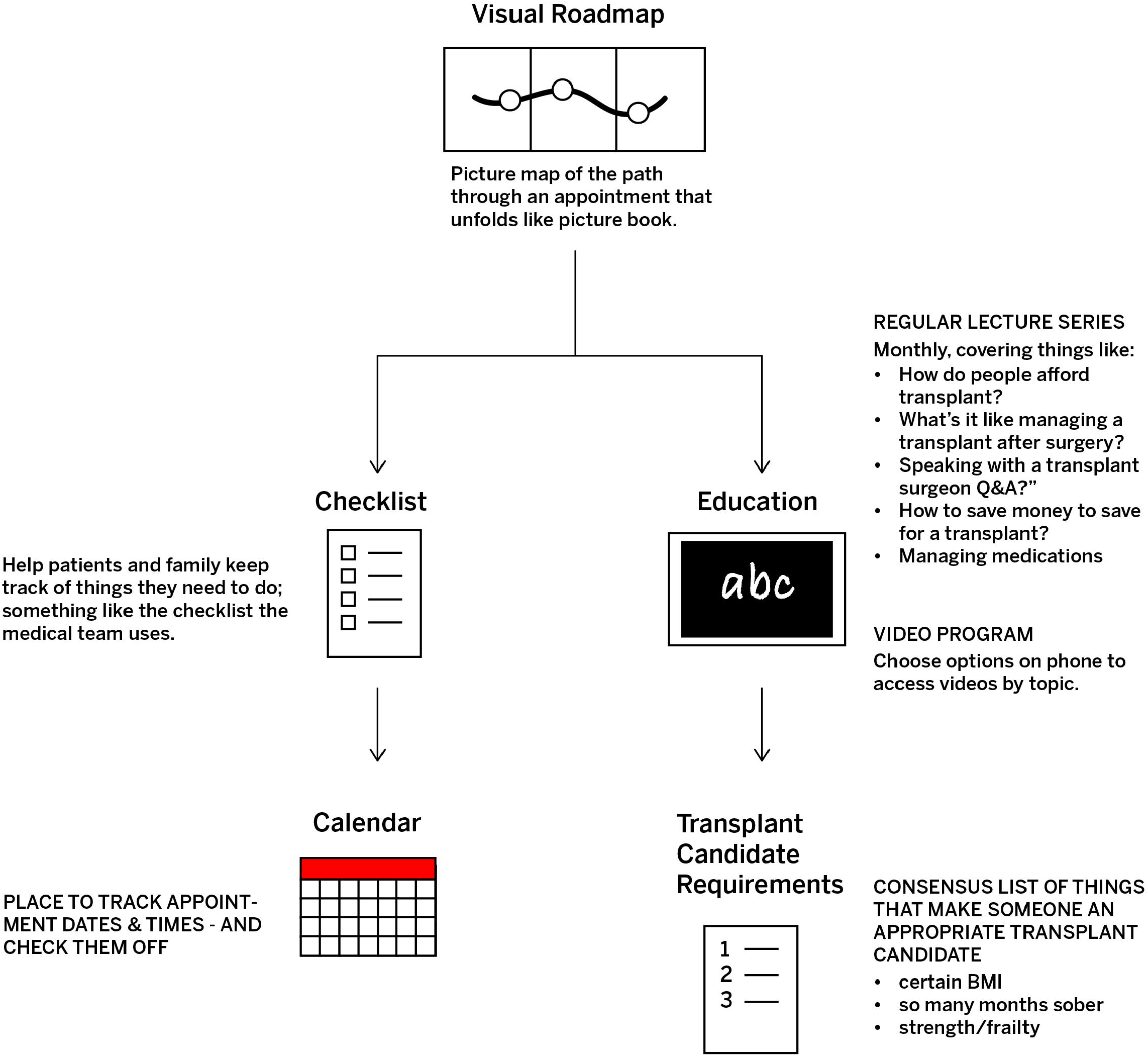
Provider-Generated Prototype:Roadmap for Transplant Education. Multimodal education roadmap to improve patient comprehension and engagement. The roadmap includes: (1) a visual map of the transplant process; (2) consensus-based criteria for transplant candidacy (e.g., sobriety, BMI, frailty); (3) a checklist and calendar to track testing and appointments; (4) a video-based education program accessible by phone; and (5) structured lecture series covering transplant expectations and post-surgical management. These components aim to reduce information overload and ensure patients and caregivers receive consistent, digestible education across milestones.

Though the providers did not create a formal prototype, they emphasized the importance of peer navigation and coordination to support patients and caregivers, connect them with resources, and guide them through HCC care. Similarly, the Transplant Evaluation Sprint, condensing pre-evaluation testing into two to three days, generated strong interest though was not formally prototyped.

After synthesis and refinement, prototypes advanced for patient feedback included a peer navigator, two Roadmap components (informational binder and patient education program), and the Evaluation Sprint (Figure 3). The Triage Pathway, while addressing evaluation challenges, required further development and had its information elements incorporated into the binder and education program to strengthen patient understanding of the LT process.

**Figure 3.**
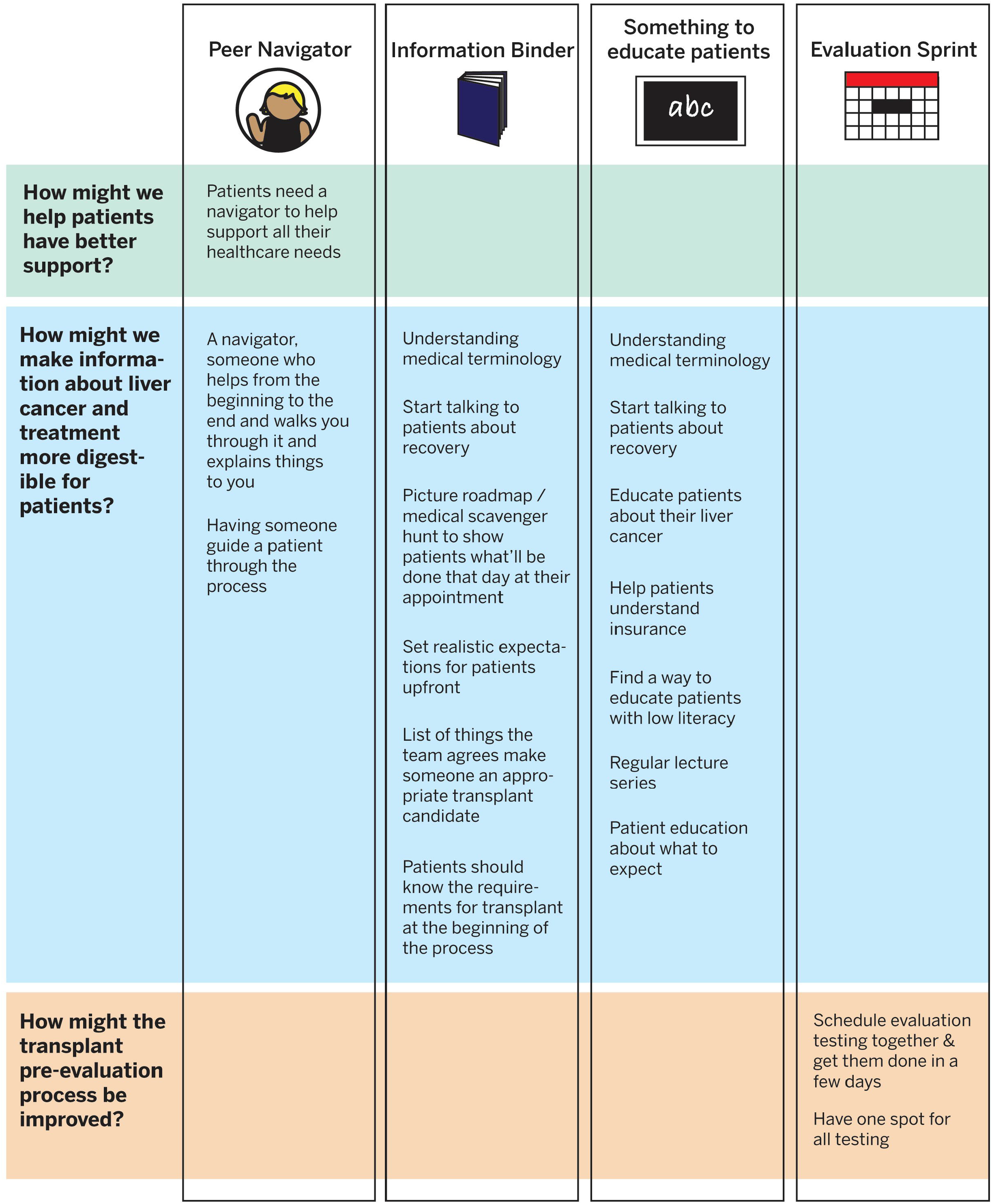
How Ideas from Participants Contributed to Final Prototypes. Final prototype concepts synthesized provider input with patient-identified barriers and solutions. Participants emphasized three core areas: streamlining evaluation (“evaluation sprint”), enhancing social support (peer navigator), and improving information delivery (education binder, multimodal education program). Ideas such as setting expectations early, simplifying medical terminology, consolidating testing, and providing ongoing recovery education informed the prototypes. Together, these concepts address inefficiencies, gaps in support, and information challenges across the transplant journey.

## DISCUSSION

This study used a stakeholder-engaged, provider-focused ideation process to generate solutions to patient-identified barriers to LT among Black and low SES patients with HCC. Through structured HCD, providers translated challenges into three intervention areas: transplant pre-evaluation inefficiencies, limited social support, and information overload. The resulting prototypes, a referral triage pathway and a transplant education roadmap, offer a scalable framework to improve care coordination, patient engagement, and equity in LT access.

Delays in LT referral and listing remain persistent barriers to equitable care, particularly for Black patients and those living in underserved communities. Inconsistent referral criteria, fragmented communication, and resource limitations frequently exacerbate these delays. The proposed triage pathway seeks to streamline pre-evaluation by introducing an early screening step, mirroring fast-track programs in oncology that have successfully shortened time to treatment and improved care continuity^15,16^. A cancer fast-track study by Martinez et al. demonstrated that structured referral pathways expedited diagnosis and therapy initiation, resulting in improved timeliness of care^15^. Similar concepts in LT could improve early identification of candidates, reduce unnecessary evaluations, and prevent system-level inefficiencies.

The triage prototype seeks to reduce inappropriate referrals and wait times through virtual pre-evaluation, standardized criteria, and improved team coordination. These efforts align with broader initiatives to enhance decision-making before referral, including clinical decision support tools for community gastroenterology practices. Together, these approaches aim to improve early identification of appropriate candidates and ensure efficient evaluation of patients most likely to benefit from transplant.

These changes are particularly important given the magnitude of disparities in LT access. Black patients with cirrhosis experience a 25% higher mortality rate and are significantly less likely to be referred or listed for LT compared to their non-Hispanic White counterparts^17^. Safety-net hospital data show that just 28% of eligible patients are referred for LT, with lower referral rates among Black patients, the uninsured, and individuals at specific hospital sites^18^. By introducing tools to improve the timing, equity, and appropriateness of referrals, this model has the potential to enhance LT equity while optimizing use of existing transplant center resources.

Information overload was a major barrier to patient understanding and engagement. In LT, patients must process complex details about treatment options and eligibility while also coping with chronic illness and cancer-related distress. For those with both HCC and cirrhosis, this burden is compounded by the need to learn about cancer staging and treatment while navigating transplant evaluation. Many seek informal sources, risking inaccurate or contradictory advice ^19^. Providers in our study echoed concerns from prior work, recommending structured, multimodal education using videos, printed guides, and digital tools^12,20^.

The education roadmap prototype is a multimodal approach that aligns with best practices in patient education, which emphasize repetition, visual reinforcement, and staged delivery to improve comprehension, recall, and engagement, particularly among patients facing complex or life-altering decisions^21^. Checklists for transplant milestones, and on-demand digital content may help reinforce key messages and reduce confusion. These tools also provide an opportunity for health systems to standardize education delivery and align messaging across care teams.

The peer navigator model builds on evidence from cancer and chronic disease management, where lay navigators improve engagement, adherence, and emotional well-being^22,23^. Navigation has been especially effective for Black and underserved patients, mitigating distress, enhancing communication, and reducing isolation^24^. Providers in this study emphasized the need for dedicated transplant navigators, ideally with lived experience, to guide patients through appointments, eligibility requirements, and post-transplant expectations. Such models have been linked to greater satisfaction, stronger therapeutic alliances, and fewer delays. Importantly, navigator programs may also reduce workload for coordinators and social workers, supporting sustainability and provider well-being.

Policy-level changes may be needed to support adoption of these interventions. Aligning navigator programs and educational efforts with CMS reimbursement structures and transplant center metrics could promote sustainability and dissemination. Integrating structured assessments of social determinants of health earlier in the evaluation may help identify at-risk patients and connect them to support.

This study has several strengths. By grounding intervention design in both patient and provider perspectives, it addresses a critical gap between disparity research and real-world implementation. The HCD framework facilitated multidisciplinary engagement and generated prototypes reflecting clinical feasibility and operational realities. Unlike top-down mandates, these interventions were co-designed with providers responsible for delivery, enhancing potential for uptake and impact.

Limitations remain. As a single-center study, some prototypes may be center-specific. While co-designed and refined with providers, direct patient testing of final prototypes is ongoing. Preliminary feedback suggests some patients prefer support groups over one-on-one navigators, which will inform adaptation. These prototypes serve as a foundation for pilot testing and multicenter evaluation, consistent with staged intervention development. Further piloting is needed to evaluate feasibility, impact on referral timelines, and outcomes such as listing rates and patient satisfaction.

The next phase will gather direct patient feedback on prototypes to ensure alignment with real-world preferences. Based on this input, the triage pathway, peer support model, and education roadmap will be revised and piloted in clinical practice. Feasibility and impact will be assessed using outcomes such as referral efficiency, time to listing, and patient-reported experience. Broader dissemination to other transplant centers will allow evaluation of scalability across diverse settings.

## CONCLUSIONS

This study shows how structured, provider-led ideation can transform patient-identified barriers into practical interventions. By focusing on triage, education, and support, these prototypes aim to improve LT access and reduce disparities. Sustained innovation, combined with stakeholder engagement and policy alignment, will be essential to translate these concepts into lasting change in LT. Importantly, these strategies may also serve as a model for advancing equity initiatives across transplant centers nationally.

## Data Availability

Interview guide available on reasonable request.

